# Tirofiban in Acute Ischemic Stroke Patients Undergoing Endovascular Thrombectomy with Preceding intravenous Thrombolysis

**DOI:** 10.1101/2023.12.07.23299704

**Authors:** Yuefei Wu, Tianrui Zhu, Jianhong Yang, Mayila Abuduaini, Feifeng Liu, Yue Zhang, Mark W. Parsons, Gang Li, Longting Lin, the INSPIRE Study Group

**Affiliations:** Shanghai East Hospital, School of Medicine, Tongji University, Shanghai, China; The First Affiliated Hospital of Ningbo University, Ningbo, Zhejiang, China; Sydney Brain Center, South West Sydney Clinical Campuses, University of New South Wales, NSW, Australia; Department of Neurology, Liverpool Hospital, New South Wales, Australia; University of Newcastle, NSW, Australia

**Keywords:** Stroke, Tirofiban, Intraarterial, Intravenous, Endovascular Thrombectomy

## Abstract

**Background:** The effectiveness of Tirofiban administration to acute ischemic stroke patients undergoing endovascular thrombectomy (EVT) after intravenous thrombolysis (IVT) remains unclear. This study examined the effect of intraarterial or intravenous tirofiban during endovascular thrombectomy following thrombolysis.

**Methods:** Patients with acute ischemic stroke who received EVT after thrombolysis were selected from the International Stroke Perfusion Imaging Registry, and divided into three groups according to tirofiban administration. Safety outcomes were symptomatic intracerebral hemorrhage (sICH) and parenchymal hematoma type-2 (PH2). Efficacy outcomes included successful recanalization, complete recanalization, functional independence, and death at 3-months. Univariate and multivariate regression estimates are listed as “estimate [95% confidence interval] p-value”.

**Results:** We analyzed a total of 682 patients who underwent EVT after IVT. Among them, 53 (7.77%) were treated with intraarterial tirofiban (IA-tirofiban group), 80 (11.73%) were treated with intravenous tirofiban (IV-tirofiban group), while 549 (80.50%) patients were not treated with tirofiban (non-tirofiban group). There were no significant differences between groups in the incidences of PH2 or sICH (*P*=0.413, *P*=0.256). There were significant differences in successful recanalization, functional independence, and death at 3-months (*P*=0.031, *P*<0.001, *P*=0.010). There was no difference between IA-tirofiban and non-tirofiban in terms of safety or efficacy outcomes. Compared with non-tirofiban, IV-tirofiban was not associated with PH2 (*P*=0.111; adjusted *P*=0.705) or sICH (*P*=0.263; adjusted *P*=0.168), but was associated with higher odds of successful recanalization (OR=8.94 [1.22–65.53], *P*=0.031; adjusted OR=8.24 [1.08–62.59], adjusted *P*=0.041), 3-month functional independence (OR=3.21 [1.88–5.50], *P*<0.001; adjusted OR=2.22 [1.21–4.12], adjusted *P*=0.011) and lower odds of 3-month death (OR=0.20 [0.17–0.27], *P*=0.007; adjusted OR=0.25 [0.07–0.92], adjusted *P*=0.039).

**Conclusions:** In acute ischemic stroke patients undergoing mechanical thrombectomy with preceding intravenous thrombolysis, both intraarterial and intravenous tirofiban could be safe. However, only intravenous tirofiban was associated with clinical benefit. Further randomized clinical trials are needed to confirm these findings.

## BACKGROUND

Endovascular thrombectomy (EVT) is becoming routine in many countries for large vessel occlusion (LVO) stroke. In patients eligible for intravenous thrombolysis (IVT), administration of tissue plasminogen activator (tPA) is recommended before the initiation of EVT. Although 71% of patients achieved successful reperfusion in previous randomized trials, only 27% of the patients treated were disability free at 90 days^1^. It is possible that either endothelial damage caused by EVT or the adverse effects of thrombolysis could activate platelet aggregation and increase thrombin activity ^2, 3^. This may lead to impaired reperfusion of the microcirculation despite complete recanalization of the artery. Thus, to improve the efficacy of reperfusion therapy, one approach is to pharmacologically prevent the aggregation of platelets.

The binding of fibrinogen to the platelet glycoprotein IIb/IIIa receptor is the final common pathway for platelet aggregation, which can be blocked by tirofiban, a nonpeptide platelet glycoprotein IIb/IIIa inhibitor^4^. Tirofiban has been proven to decrease the risk of thrombotic complications during percutaneous coronary intervention^5^. Moreover, Junghans et al^6^ indicated that tirofiban can inhibit acute thrombosis formation and reduce the likelihood of cerebral microembolism. Thus, tirofiban has been the subject of significant interest as an adjunct therapy for acute ischemic stroke (AIS). However, the use of tirofiban in AIS has been associated with mixed results. Previous studies have examined the use of tirofiban in combination either with IV-thrombolysis or EVT. There are few studies focused on tirofiban administration in patients undergoing EVT after IV-thrombolysis, and those studies that do exist focus on IA-tirofiban. In our previous study, the therapeutic effect of tirofiban during EVT in AIS was dependent on its route of administration, with intravenous tirofiban demonstrating an advantage over intraarterial tirofiban^7^. We hypothesize that this is also true in patients receiving combined IVT-EVT. Therefore, we aim to further investigate the safety and efficacy of tirofiban in AIS patients undergoing EVT after IVT.

## MATERIALS AND METHODS

### Patients

All patients were enrolled within the International Stroke Perfusion Imaging Registry (INSPIRE). The registry had central ethics approval by the Hunter New England Health District ethics committee (reference no: 11/08/17/ 4.01) and institutional ethical approvals. Written informed consent was obtained for each patient for use of their routinely collected data. This study retrospectively selected patients with acute ischemic stroke who received EVT after intravenous thrombolysis from April 2016 to September 2022. The cohort was divided into three groups according to tirofiban administration.

### EVT Procedure

All eligible patients underwent IVT followed by EVT immediately after imaging and clinical assessment of indications according to current guidelines. Anesthesia regimen (local or general), thrombectomy device and intervention strategies were at the discretion of the local interventionists and anesthetists. For cases where EVT failed to fully recanalize the occluded artery, rescue treatment with balloon angioplasty or emergency stenting was conducted.

### Tirofiban

Patient selection for tirofiban treatment (Tirofiban Hydrochloride and Sodium Chloride Injection, Grand Pharmaceutical, Co, Ltd, China) was at the discretion of the interventionalists. During the EVT procedure, the interventionists considered using tirofiban for the following indications: (1) rescue treatment with emergency stenting or balloon angioplasty for post-thrombectomy residual stenosis or failed thrombectomy; (2) prevention of distal embolization when detecting thrombus embolization likely to cause downstream arterial occlusion; (3) prevention of reocclusion when detecting intracranial atherosclerosis as the cause of LVO with a high possibility of reocclusion.

For the rescue treatment, whether a bolus injection of tirofiban was delivered intravenously or intraarterially was at the discretion of the interventionalists. For the prevention of distal embolization or reocclusion, intravenous tirofiban was more likely to be used, especially when intracranial atherosclerosis was detected. For either intraarterial tirofiban or intravenous tirofiban, the routine practice was injection a bolus dose of 10 µg/kg, followed by an intravenous infusion of tirofiban at a rate of 0.1 µg/kg/min for 12 to 24 hours. After that, intravenous tirofiban was bridged with oral antiplatelet therapy and overlapped for 4 hours before tirofiban cessation if ICH was excluded by CT 24 hours post-EVT. Based on stroke pathogenesis and head CT findings 24 hours post-EVT, oral antiplatelet (aspirin 100mg or clopidogrel 75mg once daily) or dual antiplatelet therapy were prescribed.

### Data Acquisition

We analyzed clinical characteristics, including age, baseline National Institutes of Health Stroke Scale (NIHSS), onset to arrival time, onset to IV tPA time, vascular factors, pre-stroke drug use, stroke etiology of large artery atherosclerosis (LAA), occlusion site, tirofiban administration routes and safety and efficacy outcomes. LAA was classified according to the Trial of Org 10,172 in Acute Stroke Treatment (TOAST)^8^ criteria.

### Outcomes

The safety outcomes were symptomatic intracerebral hemorrhage (sICH) and parenchymal hematoma type 2 (PH2). sICH was defined as an ICH associated with clinical deterioration according to the European Cooperative Acute Stroke Study 2 definition^9^. PH2 was assessed on non-contrast CT performed 24-hours after endovascular thrombectomy. The efficacy outcomes included successful recanalization, complete recanalization and death or disability (vs functional independence) at 3-months. Successful recanalization was defined as a modified Thrombolysis in Cerebral Ischemia (mTICI) score of 2b or 3^10^. Complete recanalization was defined as an mTICI score of 3. Functional independence was defined as a modified Rankin Scale (mRS) score of 0–2. The safety outcome and efficacy outcomes were assessed centrally and blinded to tirofiban groups.

### Statistical Analysis

Descriptive statistics were used to summarize patient characteristics and outcomes. Continuous variables and ordinal variables were described by median and interquartile range and the Kruskal-Wallis test was used to assess their differences across the 3 tirofiban groups. Categorical variables were described using percentages, with the inter-group differences assessed using the χ2 test. Univariate and multivariate regression were used to assess the effect of tirofiban group on each outcome. Confounding factors were selected based on significant differences between tirofiban groups in baseline characteristics. All statistical analysis was done using STATA 13.0 (Stata Corp, College Station, TX) with CI set at 95% and significance set at 0.05. Univariate and multivariate regression outputs are listed as “estimate [95% confidence interval], p-value”.

## RESULTS

### Patients

We analyzed a total of 682 patients who underwent EVT combined with IVT. Among them, 53 patients (7.77%) were treated with intraarterial tirofiban (IA-tirofiban group), 80 patients (11.73%) were treated with intravenous tirofiban (IV-tirofiban group), while 549 patients (80.50%) were not treated with tirofiban (non-tirofiban group). Table 1 shows the baseline characteristics of the recruited participants. Compared with the non-tirofiban group, both the IA-tirofiban and the IV-tirofiban groups demonstrated significantly higher rates of LAA (27.39%versus 73.28% versus 61.25%, *P*<0.001) and higher rates of smoking (22.39% versus 37.25% versus 28.75%, *P*=0.038). On the other hand, patients who did not receive tirofiban (non-tirofiban group) were more likely to have atrial fibrillation (AF) (48.72% versus 22.64% versus 27.50%, *P*<0.001) than patients in either the IA-tirofiban or the IV-tirofiban group. Patients in the non-tirofiban group were also older (71 versus 67 versus 67, *P*=0.010) and had higher baseline NIHSS scores (16 versus 14 versus 13, *P*<0.001). Moreover, the occlusion site and the rate of hypercholesteremia showed significant differences among the three groups (*P*=0.028, *P*=0.011, See Table 1).

**Table 1.**
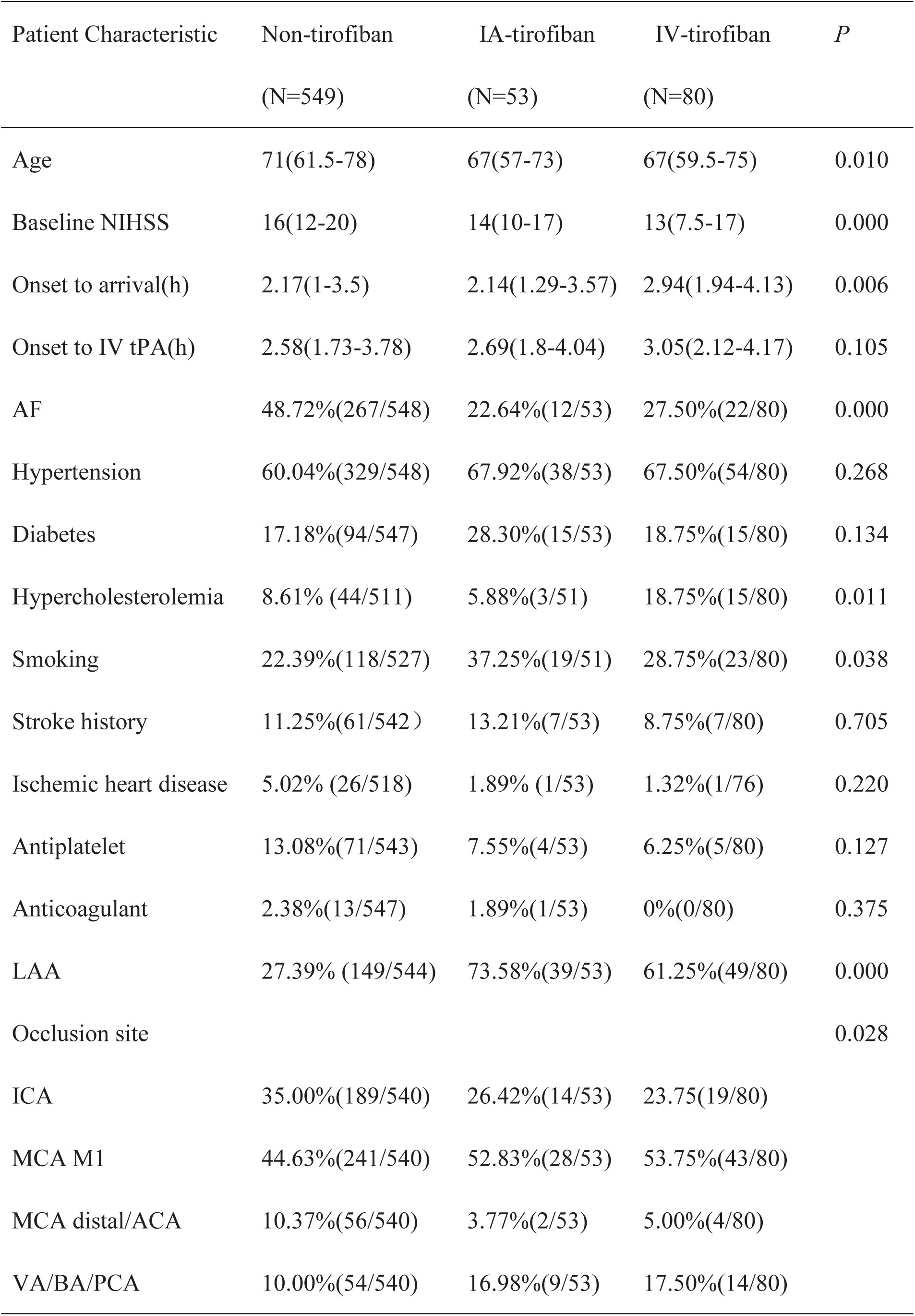
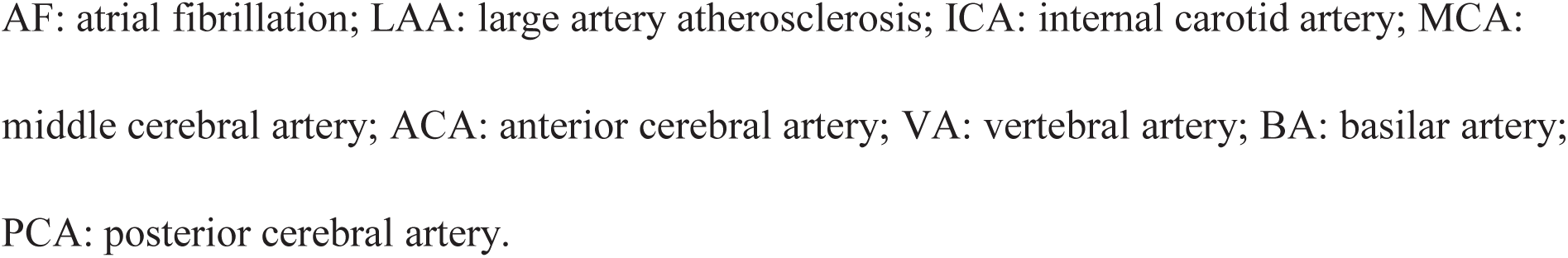
Baseline Characteristics of Patients.

| Patient Characteristic | Non-tirofiban<br>(N=549) | IA-tirofiban<br>(N=53) | IV-tirofiban<br>(N=80) | *P* |
| --- | --- | --- | --- | --- |
| Age | 71(61.5-78) | 67(57-73) | 67(59.5-75) | 0.010 |
| Baseline NIHSS | 16(12-20) | 14(10-17) | 13(7.5-17) | 0.000 |
| Onset to arrival(h) | 2.17(1-3.5) | 2.14(1.29-3.57) | 2.94(1.94-4.13) | 0.006 |
| Onset to IV tPA(h) | 2.58(1.73-3.78) | 2.69(1.8-4.04) | 3.05(2.12-4.17) | 0.105 |
| AF | 48.72%(267/548) | 22.64%(12/53) | 27.50%(22/80) | 0.000 |
| Hypertension | 60.04%(329/548) | 67.92%(38/53) | 67.50%(54/80) | 0.268 |
| Diabetes | 17.18%(94/547) | 28.30%(15/53) | 18.75%(15/80) | 0.134 |
| Hypercholesterolemia | 8.61% (44/511) | 5.88%(3/51) | 18.75%(15/80) | 0.011 |
| Smoking | 22.39%(118/527) | 37.25%(19/51) | 28.75%(23/80) | 0.038 |
| Stroke history | 11.25%(61/542) | 13.21%(7/53) | 8.75%(7/80) | 0.705 |
| Ischemic heart disease | 5.02% (26/518) | 1.89% (1/53) | 1.32%(1/76) | 0.220 |
| Antiplatelet | 13.08%(71/543) | 7.55%(4/53) | 6.25%(5/80) | 0.127 |
| Anticoagulant | 2.38%(13/547) | 1.89%(1/53) | 0%(0/80) | 0.375 |
| LAA | 27.39% (149/544) | 73.58%(39/53) | 61.25%(49/80) | 0.000 |
| Occlusion site |  |  |  | 0.028 |
| ICA | 35.00%(189/540) | 26.42%(14/53) | 23.75(19/80) |  |
| MCA M1 | 44.63%(241/540) | 52.83%(28/53) | 53.75%(43/80) |  |
| MCA distal/ACA | 10.37%(56/540) | 3.77%(2/53) | 5.00%(4/80) |  |
| VA/BA/PCA | 10.00%(54/540) | 16.98%(9/53) | 17.50%(14/80) |  |

AF: atrial fibrillation; LAA: large artery atherosclerosis; ICA: internal carotid artery; MCA: middle cerebral artery; ACA: anterior cerebral artery; VA: vertebral artery; BA: basilar artery; PCA: posterior cerebral artery.

### Safety outcomes

Raw the safety and efficacy outcomes are listed in Table 2. Occurrence of sICH was observed in 10.93%, 9.43%, and 5.00% of patients in the non-tirofiban, IA-tirofiban and IV-tirofiban groups, respectively (*P*=0.256). No significant difference in PH2 incidence was observed among the three groups either (8.74% vs. 5.66% vs. 5.00% in the non-tirofiban, IA-tirofiban and IV-tirofiban groups, *P*=0.413). Univariate regression analysis (see Table 3) found that compared to patients receiving no tirofiban, patients receiving IA-tirofiban did not experience significantly higher rates of sICH (OR= 0.85 [0.33–2.22], *P*=0.738) or PH2 (OR=0.63 [0.19–2.08], *P*=0.445). Patients receiving IV-tirofiban also demonstrated no significant increase in sICH (OR=0.43 [0.15–1.21], *P*=0.111) or PH2 (OR=0.55 [0.19–1.57], *P*=0.263) rates compared to patients receiving no tirofiban.

**Table 2.** Safety and Efficacy Outcomes.

| Patient outcomes | Non-tirofiban<br>(N=549) | IA-tirofiban<br>(N=53) | IV-tirofiban<br>(N=80) | *P* |
| --- | --- | --- | --- | --- |
| PH2 | 8.74%(48/549) | 5.66%(3/53) | 5.00% (4/80) | 0.413 |
| sICH | 10.93%(60/549) | 9.43%(5/53) | 5.00% (4/80) | 0.256 |
| Successful<br>recanalization | 89.83%(486/541) | 92.45%(49/53) | 98.75%(79/80) | 0.031 |
| Complete Recanalization | 67.84%(357/541) | 64.15%(34/53) | 80.00%(64/80) | 0.066 |
| 3-month functional<br>independence | 43.52%(215/494) | 47.06%(24/51) | 71.23%(52/73) | 0.000 |
| 3-month death | 17.81% (88/494) | 13.73% (7/51) | 4.11% (3/73) | 0.010 |

IA, intraarterial; IV, intravenous; PH2, parenchymal hematoma type-2; sICH, symptomatic intracerebral hemorrhage.

**Table 3.**
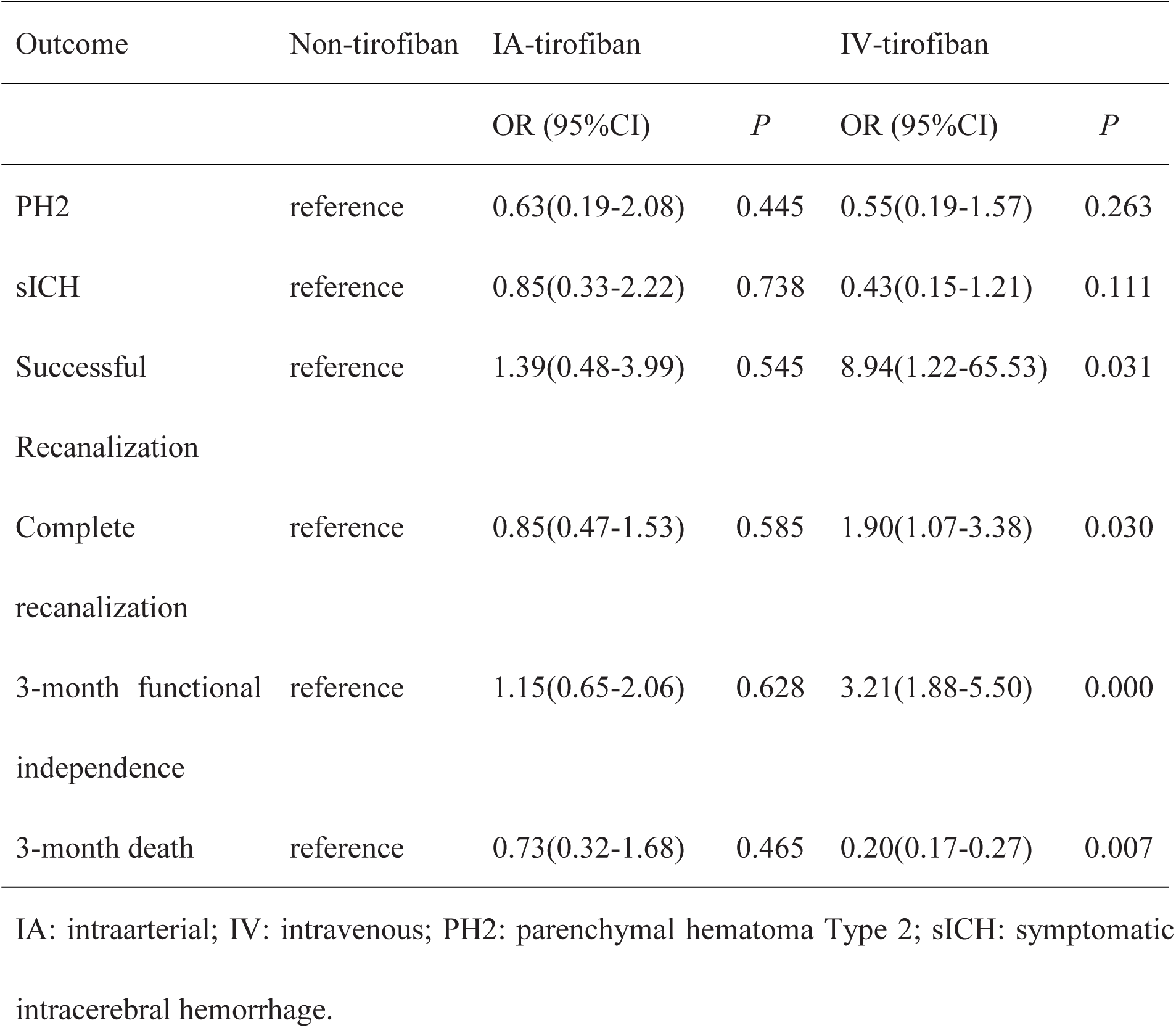
Estimated Treatment Effects of Tirofiban from Univariate Regression.

Table 4 summarizes the results of multivariate regression analysis; adjusted for smoking, hypercholesteremia, AF, LAA, age, baseline NIHSS, time to arrival, and occlusion site. Both the IA-tirofiban and the IV-tirofiban groups showed no significant difference compared to the non-tirofiban group in terms of sICH (IA-tirofiban adjusted OR=0.87 [0.28–2.64], adjusted *P*=0.805; IV-tirofiban adjusted OR=0.46 [0.16–1.38,*P*=0.168) and PH2 (IA-tirofiban adjusted OR=0.98 [0.27–3.52], adjusted *P*=0.972; IV-tirofiban adjusted OR=0.80 [0.26–2.47], adjusted *P*=0.705).

**Table 4.**
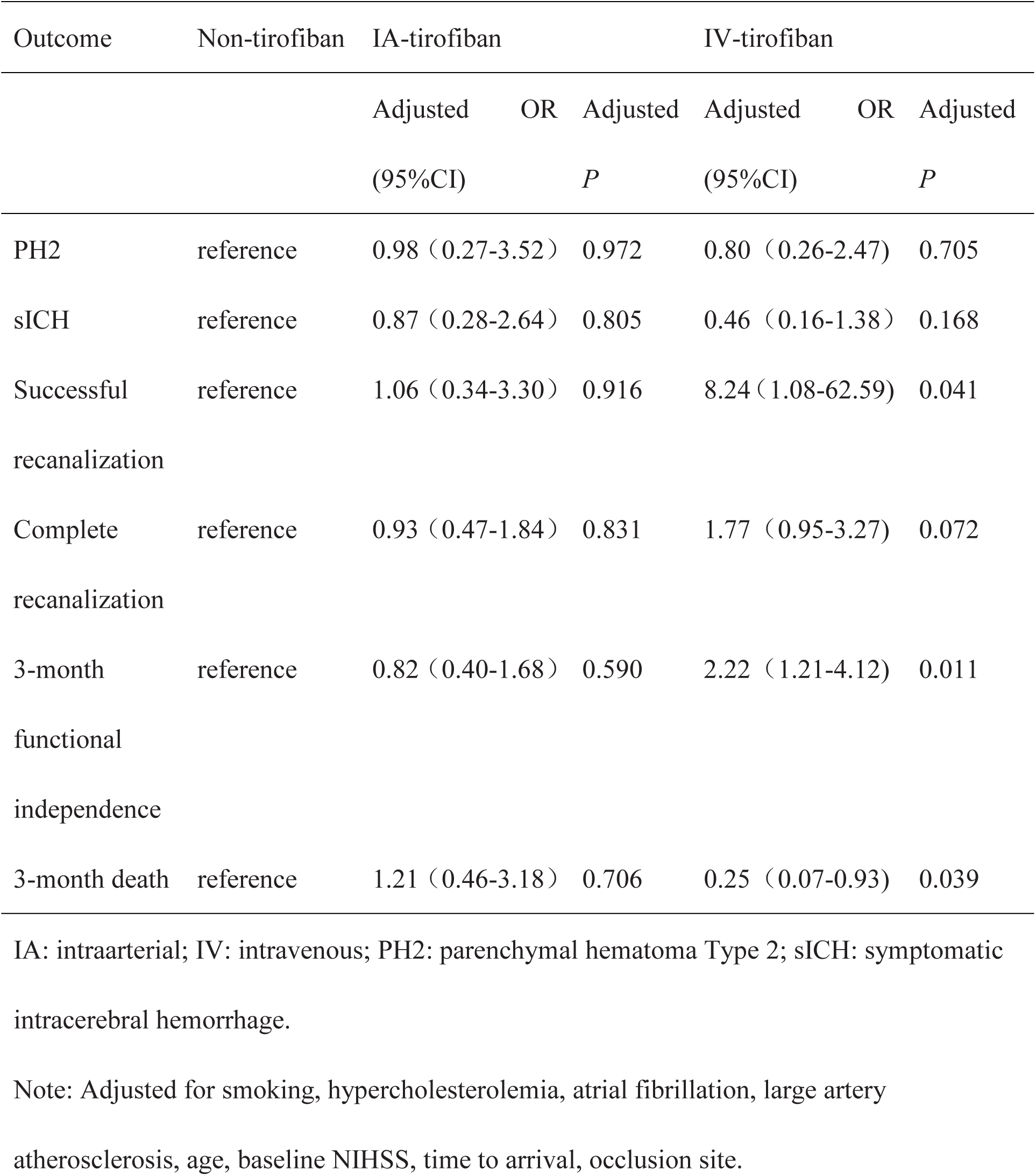
Estimated Treatment Effects of Tirofiban from Multivariate Regression.

### Efficacy outcomes

Successful recanalization (mTICI≥2b) was observed in 89.83%, 92.45%, and 98.75% of patients in the non-tirofiban, IA-tirofiban, and IV-tirofiban groups, respectively (*P*=0.031, see Table 2). Complete recanalization (mTICI=3) was observed in 67.84%, 64.15%, and 80.00% of patients in the non-tirofiban, IA-tirofiban and IV-tirofiban groups, respectively (*P*=0.066). Moreover, there were significant differences among the three groups in the rates of functional independence and death at 3 months. At 3 months, 43.52%, 47.06%, and 71.23% of patients in the non-tirofiban, IA-tirofiban and IV-tirofiban groups demonstrated functional independence (*P*<0.001). Meanwhile, death was observed in 17.81%, 13.73%, and 4.11% of patients in the non-tirofiban, IA-tirofiban and IV-tirofiban groups, respectively (*P*=0.010). Univariate regression analysis (see Table 3) showed that, compared to the non-tirofiban group, IA-tirofiban demonstrated no significant difference in terms of successful recanalization (OR=1.39 [0.48–3.99], *P*=0.545), complete recanalization (OR=0.85 [0.37–1.53], *P*=0.585), 3-month functional independence (OR=1.15 [0.65–2.06], *P*=0.628) or 3-month death (OR=0.73 [0.32–1.68], *P*=0.465). However, IV-tirofiban was independently associated with higher odds of successful recanalization (OR=8.94 [1.22-65.53], *P*= 0.031), complete recanalization (OR=1.90 [1.07–3.38], *P*=0.030), 3-month functional independence (OR=3.21 [1.88–5.50], *P*<0.001) and lower odds of 3-month death (OR=0.20 [0.17–0.27], *P*=0.007).

Multivariate regression adjusted for smoking, hypercholesteremia, AF, LAA, age, baseline NIHSS, time to arrival and occlusion site (see Table 4), showed that compared with the non-tirofiban group, IA-tirofiban was not significantly associated with successful recanalization (adjusted OR=1.06 [0.34–3.30], *P*=0.916), complete recanalization (adjusted OR=0.93 [0.47–1.84], adjusted *P*=0.831), 3-month functional independence (adjusted OR=0.82 [0.40–1.68], adjusted *P*=0.590) or 3-month death (adjusted OR=1.21 [0.46–3.18], adjusted *P*=0.706). The association between IV-tirofiban and complete recanalization did not reach statistical significance in multivariate regression (adjusted OR=1.77 [0.95–3.27], adjusted *P*=0.072). However, IV-tirofiban was independently associated with higher odds of successful recanalization (adjusted OR=8.24 [1.08–62.59], adjusted *P*=0.041), 3-month functional independence (adjusted OR=2.22 [1.21–4.12], adjusted *P*=0.011) and lower odds of 3-month death (adjusted OR=0.25 [0.07–0.92], adjusted *P*=0.039).

## DISCUSSION

Both intraarterial tirofiban and intravenous tirofiban appear to be safe in patients undergoing EVT after IVT, as assessed by the rates of sICH or PH2, but only intravenous tirofiban was associated with an increase in recanalization rates and improved clinical outcomes.

There is limited evidence available regarding optimal antiplatelet administration when performing EVT in patients treated with IV tPA. Zinkstok et al. found that early administration of aspirin after IVT was significantly associated with a higher risk of sICH in the Antiplatelet Therapy in Combination With tPA Thrombolysis in Ischemic Stroke trial^11^. Therefore, the 2018 American Heart Association/American Stroke Association guidelines indicate that aspirin administration should generally be delayed for 24 hours after the use of IVT^12^. However, early administration of tirofiban, which is a short-acting and highly selective non-peptide antagonist of the glycoprotein IIb/IIIa receptor, has been found to be safe in acute ischemic stroke patients after alteplase thrombolysis^13–15^. Therefore, tirofiban is increasingly being used in acute stroke patients undergoing mechanical thrombectomy after intravenous thrombolysis. In a study with 35 IA-tirofiban patients and 279 non-tirofiban patients, IA-tirofiban use during EVT after IVT was not associated with serious hemorrhage or 3-month mortality (adjusted OR=0.38 [0.04-1.87], *P*=0.299)^16^. One propensity-matching study with 201 patients (81 IA-tirofiban and 120 non-tirofiban) receiving EVT after IVT demonstrated no statistically significant differences in safety outcomes based on ICH, sICH, or death within 3-months, but also found no evidence of clinical benefit from IA-tirofiban (all *P*> 0.05)^17^, as did another registry study of 207 patients (55 IA-tirofiban and 152 non-tirofiban)^18^. The present study reinforces the evidence that tirofiban in patients receiving EVT after IVT is safe, and that IA-tirofiban does not confer any benefit.

To our knowledge, this is the first study assessing the safety and efficacy of IV-tirofiban in patients receiving EVT after bridging IVT. Previous studies examined IA-tirofiban in this cohort and found no evidence of efficacy in terms of recanalization or long-term functional independence. Our previous study examined the use of IV-tirofiban compared to IA-tirofiban in patients receiving EVT (most of them without bridging IVT) and found that IV-tirofiban was associated with higher rates of recanalization and functional independence compared to IA-tirofiban or no-tirofiban^7^. The reasons for the apparent advantage of IV-tirofiban compared to IA-tirofiban have yet to be elucidated but may include the following aspects. In acute stroke patients IV-tirofiban may be initiated earlier, as early as the first angiography run when intracranial atherosclerosis is detected (and before the thrombectomy procedure). This early treatment initiation allows IV-tirofiban to have a facilitating effect^19, 20^ by dissolving some platelet-rich clot before mechanical thrombectomy. Alternatively, intravenous administration is more likely to deliver tirofiban to the distal end of the clot through retrograde flow via collaterals^21^. This might help to dissolve the distal aspect of the clot and reduce the risk of reocclusion or embolism. Based on the present study and others cited above, intravenous tirofiban could be considered as an optimal antiplatelet administration for patients receiving EVT after bridging IVT.

The rates of hemorrhage in this study were considerably lower across all 3 groups (IA-tirofiban, IV-tirofiban and non-tirofiban) compared to some previous studies that examined the use of tirofiban in patients receiving EVT alone^7^. The most likely reason for this is the careful selection of patients for IVT, with clinicians often precluding the use of IVT in patients with any history or condition that would predispose them to bleeding. In addition, patients receiving bridging IVT have typically been treated within 4.5-hours of symptom onset, whereas patients may receive direct EVT up to 24-hours after onset, with an associated increase in the risk of hemorrhagic complications. Therefore, the administration of tirofiban in patients who receive bridging IVT before EVT is safe.

### Limitations

Several limitations of the present study must be taken into consideration when interpreting these results, not least the limited sample size. Specifically, only 53 patients received IA tirofiban. Importantly, this was a retrospective study without randomization. Given that the use of tirofiban was at the discretion of the neurointerventionalist, a selection bias cannot be ruled out. Specifically, the neurointerventionalists might have decided to use tirofiban only when they considered it safe based on other aspects of the patient history or clinical condition not captured in the database. However, the differences in baseline characteristics between the groups were adjusted for in multivariable regression. Moreover, this study has not recorded a prespecified initial administration time and not taken acute core volume or collateral circulation into consideration. Thus, a randomized clinical trial is still required before IV-tirofiban could be recommended as standard of care in this patient cohort.

### Conclusion

Our findings indicate that the use of tirofiban in patients who receive bridging IVT before EVT is safe. More importantly, this study suggests that IV-tirofiban should be used in the setting in preference to IA-tirofiban, and may constitute the optimum antiplatelet regimen for patients at risk of vascular reocclusion or distal embolism due to intracranial atherosclerosis.

## Data Availability

All data in the text is available

## Acknowledgments

Dr. Jianhong Yang is supported by the Traditional Chinese Medicine Science and Technology Project of Zhejiang Province (No. 2021ZA128) and Ningbo Top Medical and Health Research Program (No.2022020304)

Dr. Gang Li is supported by the Clinical Plateau Discipline Construction Project of Shanghai Pudong New Area Health Committee (No. PWYgy2021-05) and the National Natural Science Foundation of China(No. 82071192).

## References

1. Goyal M, Menon BK, van Zwam WH, Dippel DW, Mitchell PJ, Demchuk AM, et al. Endovascular thrombectomy after large-vessel ischaemic stroke: A meta-analysis of individual patient data from five randomised trials. Lancet. 2016;387:1723–1731.doi: 10.1016/S0140-6736(16)00163-X.

2. Power S, Matouk C, Casaubon LK, Silver FL, Krings T, Mikulis DJ, et al. Vessel wall magnetic resonance imaging in acute ischemic stroke: Effects of embolism and mechanical thrombectomy on the arterial wall. Stroke,2014;45:2330–2334. doi:10.1161/STROKEAHA.114.005618.

3. Saqqur M, Molina CA, Salam A, Siddiqui M, Ribo M, Uchino K, et al. Clinical deterioration after intravenous recombinant tissue plasminogen activator treatment: A multicenter transcranial doppler study. Stroke. 2007;38:69–74. doi:10.1161/01.STR.0000251800.01964.f6.

4. Lefkovits J, Plow Ef Fau - Topol EJ, Topol EJ. Platelet glycoprotein iib/iiia receptors in cardiovascular medicine. N Engl J Med. 1995;332:1553–1559. doi:10.1056/NEJM199506083322306.

5. Cannon CP, Weintraub WS, Demopoulos LA, Vicari R, Frey MJ, Lakkis N, et al. Comparison of early invasive and conservative strategies in patients with unstable coronary syndromes treated with the glycoprotein iib/iiia inhibitor tirofiban. N Engl J Med. 2001;345:1879–1887. doi:10.1056/NEJM200106213442501.

6. Junghans, U. Cerebral microembolism is blocked by tirofiban, a selective nonpeptide platelet glycoprotein iib/iiia receptor antagonist. Circulation.2003;107:2717–2721.doi: 10.1161/01.CIR.0000070544.15890.0E.

7. Yang J, Wu Y, Gao X, Bivard A, Levi CR, Parsons MW, et al. Intraarterial versus intravenous tirofiban as an adjunct to endovascular thrombectomy for acute ischemic stroke. Stroke. 2020;51:2925–2933.doi:10.1161/STROKEAHA.120.029994.

8. Adams HP, Bendixen BH, Kappelle LJ, Biller J, Love BB, Gordon DL, et al. Classification of subtype of acute ischemic stroke. Definitions for use in a multicenter clinical trial. Toast. Trial of org 10172 in acute stroke treatment. Stroke. 1993;24:35–41. doi:10.1161/01.str.24.1.35.

9. Hacke W, Kaste M, Fieschi C, von Kummer R, Davalos A, Meier D, et al. Randomised double-blind placebo-controlled trial of thrombolytic therapy with intravenous alteplase in acute ischaemic stroke (ecass ii). Second european-australasian acute stroke study investigators. Lancet. 1998;352:1245–1251. doi:10.1016/s0140-6736(98)08020-9.

10. Zaidat OO, Yoo AJ, Khatri P, Tomsick TA, Rüdiger von K, Saver JL, et al. Recommendations on angiographic revascularization grading standards for acute ischemic stroke: A consensus statement. Stroke. 2013;44:2650–2663. doi:10.1161/STROKEAHA.113.001972.

11. Zinkstok SM, Roos YB, ARTIS investigators. Early administration of aspirin in patients treated with alteplase for acute ischaemic stroke: A randomised controlled trial. Lancet. 2012;380:731–737.doi:10.1016/S0140-6736(12)60949-0.

12. Powers WJ, Rabinstein AA, Ackerson T, Adeoye OM, Bambakidis NC, Becker K, et al. 2018 guidelines for the early management of patients with acute ischemic stroke: A guideline for healthcare professionals from the american heart association/american stroke association. Stroke. 2018;49:e46–e110.doi:10.1161/STR.0000000000000158.

13. Li W, Lin L, Zhang M, Wu Y, Liu C, Li X, et al. Safety and preliminary efficacy of early tirofiban treatment after alteplase in acute ischemic stroke patients. Stroke. 2016;47:2649–2651.doi:10.1161/STROKEAHA.116.014413.

14. Wu C, Sun C, Wang L, Lian Y, Xie N, Huang S, et al. Low-dose tirofiban treatment improves neurological deterioration outcome after intravenous thrombolysis. Stroke. 2019;50:3481–3487.doi:10.1161/STROKEAHA.119.026240.

15. Li W, Lin G, Xiao Z, Zhang Y, Li B, Zhou Y, et al. Safety and efficacy of tirofiban during intravenous thrombolysis bridging to mechanical thrombectomy for acute ischemic stroke patients: A meta-analysis. Frontiers in Neurology. 2022;13. doi:10.3389/fneur.2022.851910.

16. Jang SH, Sohn SI, Park H, Lee SJ, Kim YW, Hong JM, et al. The safety of intra-arterial tirofiban during endovascular therapy after intravenous thrombolysis.Am J Neuroradiol. 2021;42:1633–1637.doi:10.3174/ajnr.A7203.

17. Ma G, Li S, Jia B, Mo D, Ma N, Gao F, et al. Safety and efficacy of low-dose tirofiban combined with intravenous thrombolysis and mechanical thrombectomy in acute ischemic stroke: A matched-control analysis from a nationwide registry. Frontiers in neurology. 2021;12:666919.doi:10.3389/fneur.2021.666919.

18. Huo X, Yang M, Ma N, Gao F, Mo D, Li X, et al. Safety and efficacy of tirofiban during mechanical thrombectomy for stroke patients with preceding intravenous thrombolysis. Clin Interv Aging. 2020;15:1241–1248.doi:10.2147/CIA.S238769.

19. Liu J, Shi Q, Sun Y, He J, Yang B, Zhang C, et al. Efficacy of tirofiban administered at different time points after intravenous thrombolytic therapy with alteplase in patients with acute ischemic stroke. J Stroke Cerebrovasc Dis. 2019;28:1126–1132.doi: 10.1016/j.jstrokecerebrovasdis.2018.12.044.

20. Montalescot G, Borentain M, Payot L, Collet JP, Thomas D. Early vs late administration of glycoprotein iib/iiia inhibitors in primary percutaneous coronary intervention of acute st-segment elevation myocardial infarction: A meta-analysis. Acc Current Journal Review. 2004;13:30–31.doi:10.1001/jama.292.3.362.

21. Bang OY, Saver JL, Kim SJ, Kim GM, Chung CS, Ovbiagele B, et al. Collateral flow predicts response to endovascular therapy for acute ischemic stroke. Stroke. 2011;42:693–699.doi: 10.1161/STROKEAHA.110.595256.

